# Stage 2a IDEAL evaluation of a third-generation biocomposite suture anchor in arthroscopic rotator cuff repair: Subgroup cohort analysis of the PRULO registry with 12-month follow up

**DOI:** 10.1101/2024.06.13.24307996

**Authors:** Corey Scholes, Manaal Fatima, Cooper Moody, Kevin Eng, Richard S Page

**Affiliations:** EBM Analytics, Sydney, New South Wales, Australia; Barwon Health, Geelong, Victoria, Australia; Geelong Orthopaedics, Geelong, Victoria, Australia; Barwon Centre for Orthopaedic Research and Education (B-CORE), Deakin University, Victoria, Australia; St John of God Hospital, Geelong, Victoria, Australia

**Author notes:** **Corresponding author** Richard S Page Geelong Orthopaedics Level 2/83 Myers St, Geelong VIC 3220, Australia.

**Keywords:** Rotator cuff repair, biocomposite, Poly-lactic co-glycolic acid, tricalcium phosphate, Patient-reported outcomes, IDEAL

## Abstract

**Background:** Poly-lactic co-glycolide with tricalcium phosphate (PLGA)/β-TCP is both bioactive and biodegradable, and is considered a third generation biomaterial for suture anchors. This study aims to describe the incidence of adverse events and the trajectory of patient-reported outcomes up to 12 months follow up in patients undergoing rotator cuff repair with this type of anchor.

**Methods:** A sub-group analysis of a prospective clinical registry embedded in an orthopaedic clinic was conducted. Patients undergoing surgery with the Healix Advance BR were identified and data on patient demographics, treatment details, complications and adverse events, as well as patient-reported outcomes (QuickDASH, WORC Index Normalised) were retrieved. Summary statistics were generated for patient characteristics and PROMs analysed using multiple imputation and a linear model to assess changes between baseline and 12 month follow up.

**Results:** A consecutive cohort of 68 cases receiving the anchor of interest was included for analysis. Complications were recorded in six cases (8.8%, 95%CI 3.6 - 18.9), presenting as postoperative capsulitis/stiffness (N=3), persistent shoulder pain (N=1), a case of hemi-diaphragm palsy and a case of complex regional pain syndrome in the hand. No infections or reoperations were encountered in this series. The QuickDASH scores improved significantly from 45 (IQR 34-57) preoperatively to 5 (IQR 2-22) at 12 months. WORC Index scores improved from 46 (IQR 27-60) preoperatively to 85 (IQR 62-93) at 12 months.

**Conclusion:** This study found a low incidence of adverse events, no requirement for reoperation or revision, and favourable recovery of patient-reported outcomes in patients treated with arthroscopic rotator cuff repair with a third-generation biocomposite suture anchor at up to 12 months follow up.

## Introduction

Symptomatic tears of the rotator cuff remain a prevalent condition (Rees, 2008) and arthroscopic surgical repair is undertaken with increasing annual incidence (Colvin et al., 2012). A high risk of subsequent retear and revision arthroscopic repair persist despite advances in materials and techniques. In conventional suture anchors used for primary surgery, the incidence of revision procedures has been estimated at up to 27% (estimate 20%; 95%CI 15.3 - 26.8) (Boksh et al., 2022).

Biostable suture anchors remain popular for rotator cuff repair fixation, but have been associated with adverse events such as loosening, chondral damage, as well as interfering with revision procedures and post-operative imaging (Suroto et al., 2023). Biocomposite anchors offer advantages in rotator cuff surgery with reduced risk of suture damage in-vivo (Savage et al., 2023; Yang et al., 2023) and the replacement of bioresorbable anchors with native bone, thereby restoring anatomy (Barber et al., 2013). The absence of hardware to remove and the restoration of bone stock may reduce the complexity of subsequent revision procedures.

The concept of biodegradable anchors is well established, however there may be a role for new materials and designs to reduce cyst formation incidence (Kim et al., 2015) and improve bone integration. The results of suture anchors comprising one specific material for cuff repair is not easily discerned from the existing literature, with biodegradable materials usually combined in review papers. One systematic review (Barber et al., 2017) estimated resorption rates of at least 60% at 30 month follow up, with a low rate of imaging-detected complications. A more recent systematic review (Suroto et al., 2023) reported tendon healing rates ranging from 50 - 100% in four studies of biodegradable anchors in rotator cuff repair (RCR).

Poly-lactic co-glycolic acid-coated tricalcium phosphate (PLGA)/β-TCP) is a biocomposite material explicitly developed to promote absorption at a controlled rate (Cho et al., 2021), and being both bioactive and biodegradable is considered a 3rd-generation biomaterial for suture anchors (Filip et al., 2022). A specific example is the Healix Advance BR (Depuy-Mitek, USA; Biocryl Rapide, 30% b-tricalcium phosphate [β-TCP]/70% poly [lactide co-glycolide] [PLGA]), which is a screw-in anchor with a distal bar eyelet and can be double or triple loaded with No.2 suture (Barber & Herbert, 2013). It has shown satisfactory ultimate failure load during in-vitro testing (Patzer et al., 2011), but lower failure loads during cyclic loading in both cortical and cancellous bone compared to contemporary designs comprising different materials (Barber & Herbert, 2013). Nevertheless, pain reduction and recovery of cyst formation over time has been demonstrated, with a reported retear rate of 13% at 12 month follow up (Chung et al., 2018). Equivalent patient-reported outcomes have also been reported between this suture anchor and all-suture design at up to 8 months, (Di Gennaro et al., 2022).

The information available on this anchor is encouraging but considering the IDEAL framework of surgical innovation (McCulloch et al., 2013), the evaluation of its clinical performance remains in the early stages of *idea* and *development*. Therefore, it is imperative that evidence is generated around the clinical performance of implants of this type and material. In particular, assessment in diverse populations is a priority. Institutional registries (Orthopaedics1*, 2019) offer a relatively robust framework for evaluation of innovative treatments and technologies within a variety of healthcare settings and populations (Gliklich et al., 2019). The capacity to prospectively enrol patients and standardise follow up alleviates some methodological weaknesses of retrospective observational studies. Our group has emulated a quality registry in clinical practice (Scholes et al., 2023) which provides a unique platform for contributing to the evidence base of this biodegradable anchor in a diverse population.

This paper presents a Stage 2a IDEAL (McCulloch et al., 2013) evaluation of a third-generation biocomposite suture anchor in a registry sub-analysis of patients presenting with repairable rotator cuff tear for arthroscopic repair in a regional practice, describing the incidence of all-cause failures, adverse events and the trajectory of patient-reported outcomes up to 12 months follow up.

## Methods

### Study Design

Retrospective sub-group analysis of a prospective clinical registry embedded in a regional, orthopaedic clinic. The RECORD guidelines (Benchimol et al., 2015) were observed in the reporting of this study (Supplementary 1).

### Setting

Data was retrieved from a multi-cohort, prospective observational, clinical quality registry. The Patient Registry of Upper Limb pathology Outcomes (PRULO) is a single-centre study involving three clinician investigators, collating clinical data and patient-reported outcomes for patients presenting to a specialist orthopaedic clinic with upper limb pathology (Scholes et al., 2023).

### Registration and Ethics

Ethical approval for PRULO was obtained from the Barwon Health Research Ethics Committee (Project ID 49184, Reference 19/70), and the registry is listed on the Australian New Zealand Clinical Trials Registry (ACTRN12619000770167).

### Patient Selection

Patients were included in the PRULO registry as per the registry inclusion and exclusion criteria (Scholes et al., 2023). A cohort of patients undergoing rotator cuff repair surgery with the anchor material of interest were identified by cross-matching product stock keeping units (SKU), with packaging information retained in surgery (Supplementary 2). Patients were included in the analysis irrespective of patient or tear characteristics.

### Surgical Technique and Perioperative Management

The present single-surgeon series involved an arthroscopic rotator cuff repair (ARCR), performed with double row transosseous equivalent repairs in the beach chair position. The anchor of interest (Healix Advance BR, Depuy-Mitek) was utilised with a self-tensioning suture (Dynacord or Orthocord, Depuy-Mitek) or tape (Permatape, Depuy-Mitek) and free suture was used when margin convergence was performed. Adjunct procedures included tenodesis or tenotomy of the long head biceps proximal tendon, limited subacromial decompression and lateral clavicle resection in some cases. Postoperative recovery involved shoulder immobilisation for four weeks and elbow and hand physiotherapy from day one. Shoulder motion was restricted to passive assisted for the next four weeks, and increased to active assisted range of motion from 8-12 weeks, with isometric exercises at 8 weeks and strengthening exercises added at 12 weeks.

### Outcomes

Variables collected in the PRULO registry span patient data, clinical and treatment data, and outcomes data including patient-reported outcome measures (Scholes et al., 2023). Variables retrieved to address the study aims (see Supplementary 2) included:

● Patient demographics (age, sex, body mass index (BMI), hand dominance)
● Treatment details (bilateral procedures, tear characteristics)
● Complications and adverse events. The IDEAL framework identifies scope and severity of complications as common items for which agreed standard definitions are required across the international surgical community (McCulloch et al., 2013). Since then, substantial work has been conducted to standardise complication reporting after ARCR (Audigé et al., 2016) and a modification of the Sink severity grading (Sink et al., 2012) specific for rotator cuff surgery (Felsch et al., 2021) was used to grade each instance of complication.
● Procedure survival, with treatment failure classified as a revision of the original procedure
● Patient-reported outcomes (Quick Disabilities of the Arm, Shoulder and Hand (QuickDASH) questionnaire and the Western Ontario Rotator Cuff (WORC) Index)

○ The QuickDASH was calculated according to the developer’s user manual (Institute for Work & Health, n.d.)
○ The WORC Index (Normalised) was calculated according to (Kirkley et al., 2003).
○ One question response (Physical Q3 - *How much weakness do you experience in your shoulder?*) was analysed in isolation.

### Statistical Analysis

Descriptives for continuous variables of patient characteristics were reported using medians and interquartile ranges, counts and proportions were reported categorical variables. If multiple complications were retrieved for each treatment, they were recorded when they presented and descriptive statistics generated for all complications observed. For procedure survival, duration (surgery date to *enddate*) was summarised with mean and standard deviation and visualised with ridge plots using a distribution plotting package *ggdist* (Kay, 2023). A Kaplan-Meier survival curve (with 95% confidence intervals) with procedure *failure* as the event of interest was created with the *survival* package and plotted using *ggsurvfit*. A linear model was applied to each imputed dataset for QuickDASH and WORC Index, with *Timepoint* as the primary predictor and *Age at surgery* and *Sex* included as covariates. The results from each model were pooled in the *gtsummary* package for presentation including coefficients, 95% confidence intervals and p-values. P-values for Sex and Timepoint were reported for comparisons against a reference level. The *marginaleffects* package was used to calculate pooled predictions for the PROMs based on the linear models across the multiple-imputed datasets (Arel-Bundock, 2023) and plotted across *Timepoint*. Further details can be found in Supplementary 2.

## Results

### Patient characteristics

A sample of 916 cases electing to undergo surgery within the registry were assessed for eligibility, with 68 receiving surgery with the anchor of interest included in the final analysis. There were no withdrawals of consent from the registry, one case was excluded from PROMs followup due to enrollment in an external randomised trial. Compliance with PROMs was 51.7% at 12 months for QDASH and 43.1% for WORC (Supplementary 2). The analysis cohort comprised 31% females, had a mean age of 61 years (IQR 58-63) and a mean body mass index (BMI) of 23 (IQR 18-29). Surgery was performed on the dominant side for 60% of patients, and 16% of the cohort underwent sequential bilateral procedures. The majority of patients (98%) presented with a full tear (Table 2).

**Table 1:**
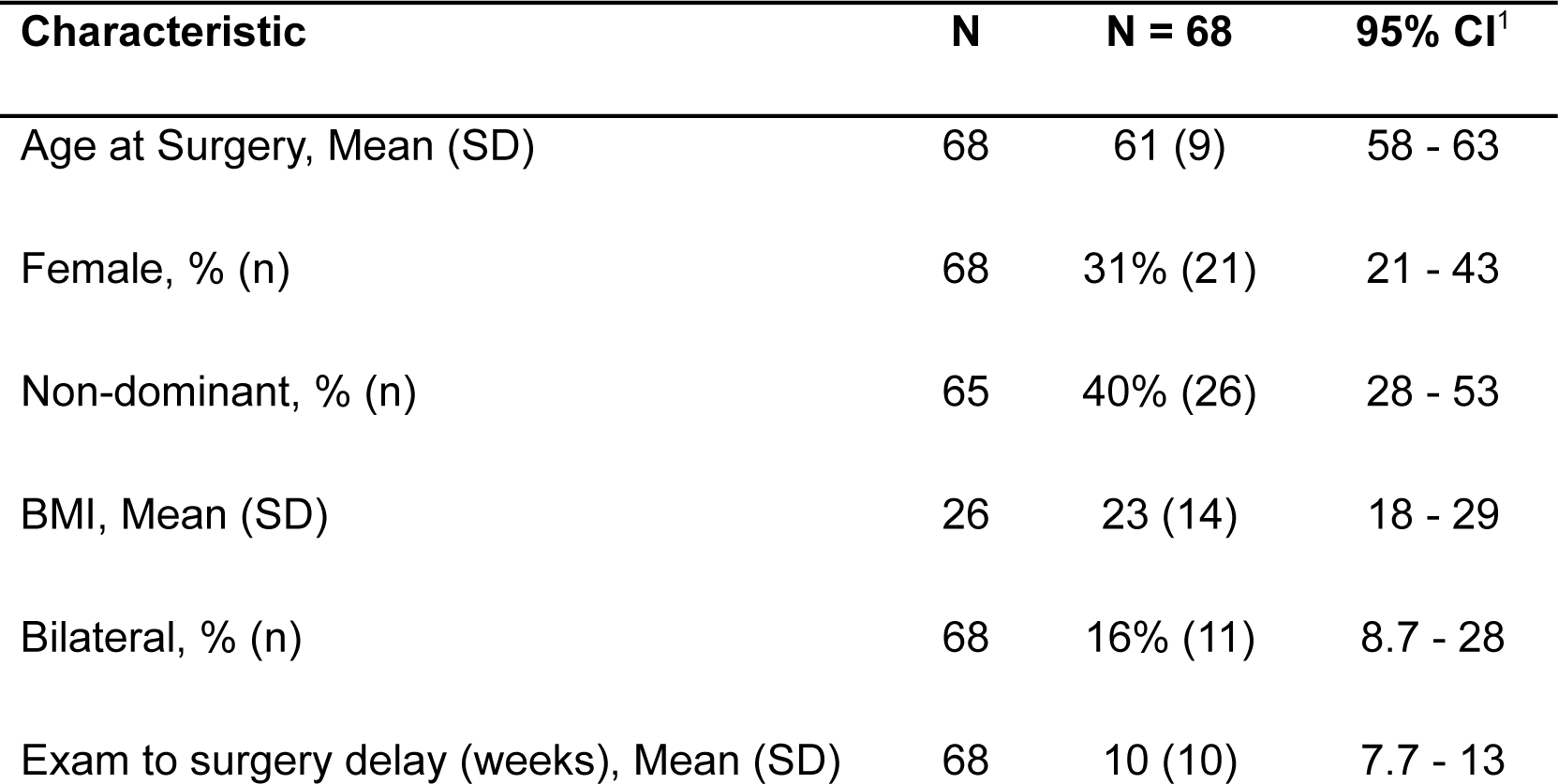
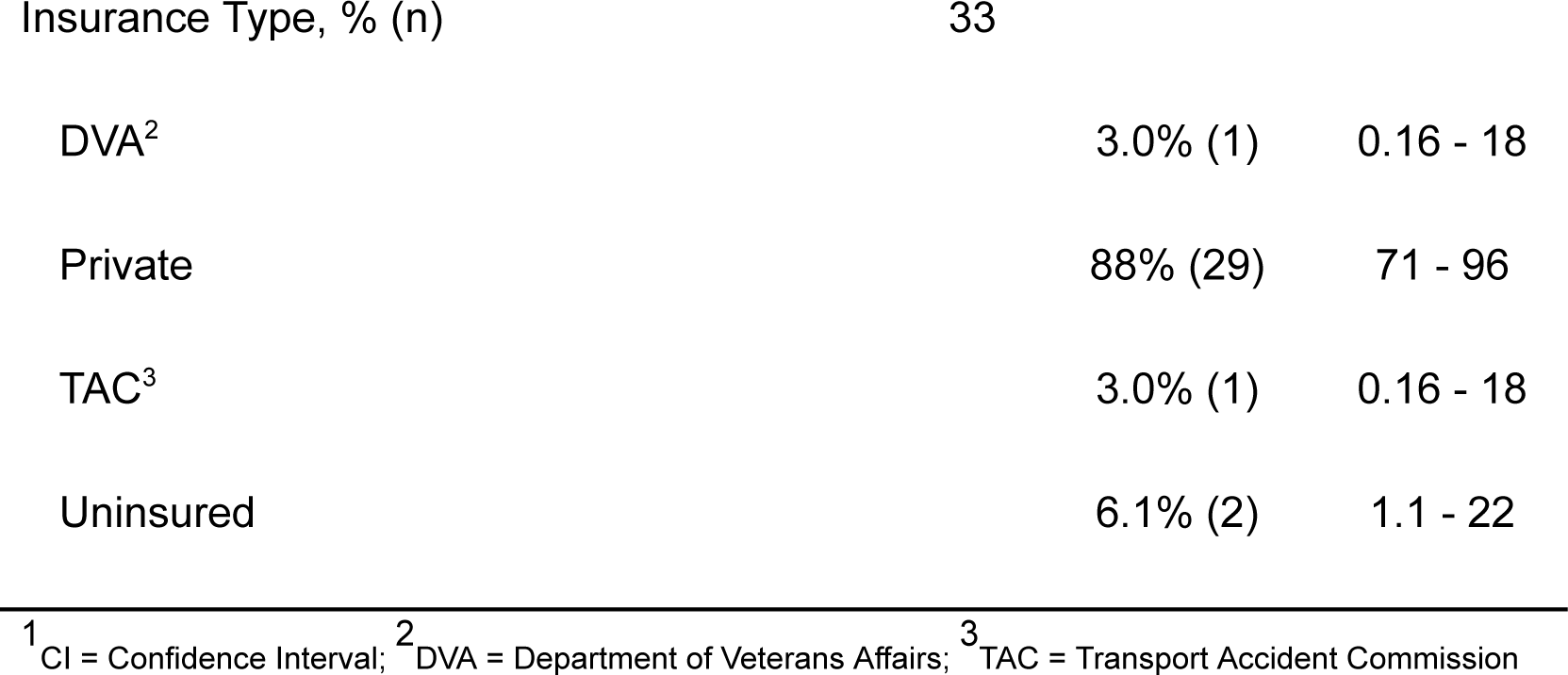
Summary of pre-operative patient characteristics for the cohort.

**Table 2:** Summary of pathology characteristics for the cohort.

| Characteristic | Available<br>Sample | Summary<br>Statistic | 95% CI<sup>1</sup> |
| --- | --- | --- | --- |
| Primary Presentation, % (n) | 68 | 100 (68) | 93 - 100 |
| Full Tear, % (n) | 63 | 98 (62) | 90 - 100 |
| Fatty Infiltration, % (n)<sup>2</sup> | 63 |  |  |
| 0 |  | 11 (7) | 5.0 - 22 |
| 1 |  | 51 (32) | 38 - 63 |
| 2 |  | 33 (21) | 22 - 46 |
| 3 |  | 4.8 (3) | 1.2 - 14 |
| Tendon Retraction, % (n)<sup>3</sup> | 63 |  |  |
| I |  | 33 (21) | 22 - 46 |
| II |  | 44 (28) | 32 - 57 |
| III |  | 19 (12) | 11 - 31 |
| IV |  | 3.2 (2) | 0.55 - 12 |
| Tendon Delamination, % (n) | 63 | 71 (45) | 58 - 82 |
| Tendons Involved (+Supraspinatus), % (n) | 63 |  |  |
| Infraspinatus |  | 22 (14) | 13 - 35 |
| Infraspinatus; Subscapularis |  | 11 (7) | 5.0 - 22 |
| Infraspinatus; Teres Minor; Subscapularis |  | 1.6 (1) | 0.08 - 9.7 |
| Subscapularis |  | 13 (8) | 6.0 - 24 |
| Subscapularis (isolated) |  | 16 (10) | 8.3 - 28 |
| Supraspinatus (isolated) |  | 37 (23) | 25 - 50 |
| Tear Size AP (mm), Mean (SD) | 62 | 24 (11) | 21 - 27 |
| Tear Size ML (mm), Mean (SD) | 63 | 19 (9) | 17 - 22 |
| Tear Area (mm<sup>2</sup>), Mean (SD) | 62 | 551 (588) | 402 - 700 |
| Tear Classification, % (n)<sup>4</sup> | 62 |  |  |
| Large |  | 16 (10) | 8.4 - 28 |
| Massive |  | 3.2 (2) | 0.56 - 12 |
| Medium |  | 76 (47) | 63 - 85 |
| Partial |  | 1.6 (1) | 0.08 - 9.8 |
| Small |  | 3.2 (2) | 0.56 - 12 |
| Tear Pattern, % (n) | 63 |  |  |
| Crescent |  | 49 (31) | 37 - 62 |
| L |  | 17 (11) | 9.4 - 30 |
| Reverse L |  | 14 (9) | 7.1 - 26 |
| U |  | 16 (10) | 8.3 - 28 |
| V |  | 3.2 (2) | 0.55 - 12 |
| Other Pathology, % (n) | 46 | 43 (20) | 29 - 59 |

<sup>1</sup>CI = Confidence Interval; <sup>2</sup>Fuchs et al 1999; <sup>3</sup>Modified Patte Grading (Lädermann et al., 2016);
<sup>4</sup>(Rashid et al., 2017)

### Complications and Treatment Survival

There were no intraoperative adverse events, or any instances of explantation of the anchor. Postoperative complications were reported in six cases (overall incidence of 8.8%, 95%CI 3.6 - 18.9). Three cases presented with capsulitis - progressive pain and stiffness, at 8-21 weeks after surgery (Sink grade I) and one receiving a corticosteroid injection (Sink grade II). One of these cases displayed signs of adhesive capsulitis at the time of surgery. One case suffered from transient phrenic nerve palsy suspected to be due to a local anaesthetic interscalene block prior to the repair procedure (Sink grade II). One case re-presented with complex regional pain syndrome in the ipsilateral limb at 16 weeks follow-up, while one case presented with persistent post-surgical pain at 16 weeks follow-up (Sink grade I). No infections or inflammatory reactions were observed in this series. No reoperation procedures were performed during the duration of follow-up, and no treatment failures were observed.

### Patient-reported outcomes

The model-predicted QuickDASH improved significantly from baseline to 12 month follow up (Table 3). QuickDASH scores for the cohort were 45 (IQR 34-57) preoperatively, 32 (IQR 20-48) at three months, 18 (IQR 11-30) at six months and 5 (IQR 2-22) at 12 months. Average scores improved over the 12 months following surgery, and the distribution of scores revealed different trajectories of recovery for the patients, with the majority responding well to treatment (Figure 2).

**Table 3:** Summary of model of QuickDASH after multiple imputation.

| Characteristic | Beta | 95% CI<sup>1</sup> | p-value |
| --- | --- | --- | --- |
| TimePoint |  |  |  |
| Preop | — | — |  |
| 3 months | -12.11 | -18.28, -5.95 | <0.001 |
| 6 months | -25.38 | -32.11, -18.65 | <0.001 |
| 12 months | -32.11 | -38.50, -25.71 | <0.001 |
| Age at Surgery | -0.03 | -0.43, 0.37 | 0.873 |
| Male vs Female | -8.19 | -16.39, 0.00 | 0.050 |

<sup>1</sup>CI = Confidence Interval

Table 3: Summary of models of WORC Index and Physical Q3 after multiple imputation
| Characteristic | Normalised Index |  |  | Physical Q3 |  |  |
| --- | --- | --- | --- | --- | --- | --- |
|  | Beta | 95% CI<sup>1</sup> | p-value | Beta | 95% CI<sup>1</sup> | p-value |
| TimePoint |  |  |  |  |  |  |
| Preop | — | — |  | — | — |  |
| 6 months | 31.81 | 18.54, 45.08 | <0.001 | -30.56 | -43.94, -17.18 | <0.001 |
| 12 months | 38.69 | 28.21, 49.16 | <0.001 | -43.75 | -55.59, -31.91 | <0.001 |
| Age at Surgery | 0.28 | -0.18, 0.74 | 0.229 | -0.43 | -1.13, 0.26 | 0.204 |
| Male vs Female | 9.18 | -1.11, 19.47 | 0.078 | -8.84 | -21.47, 3.79 | 0.161 |

<sup>1</sup>CI = Confidence Interval

Timepoint was associated with significant improvements in the WORC Index from baseline when adjusted for age and sex (Table 3). WORC Index scores for the cohort were 46 (IQR 27-60) preoperatively, 74 (IQR 57-87) at six months and 85 (IQR 62-93) at 12 months. As with the QuickDASH, scores improved over the 12 months following surgery, and the distribution of scores revealed different trajectories of recovery for the patients (Figure 1). The individual question (Physical Q3) demonstrated improvements in the sample average over time, but a proportion reported persistent weakness in the shoulder at the 12 months after surgery (Figure 3).

**Figure 1:**
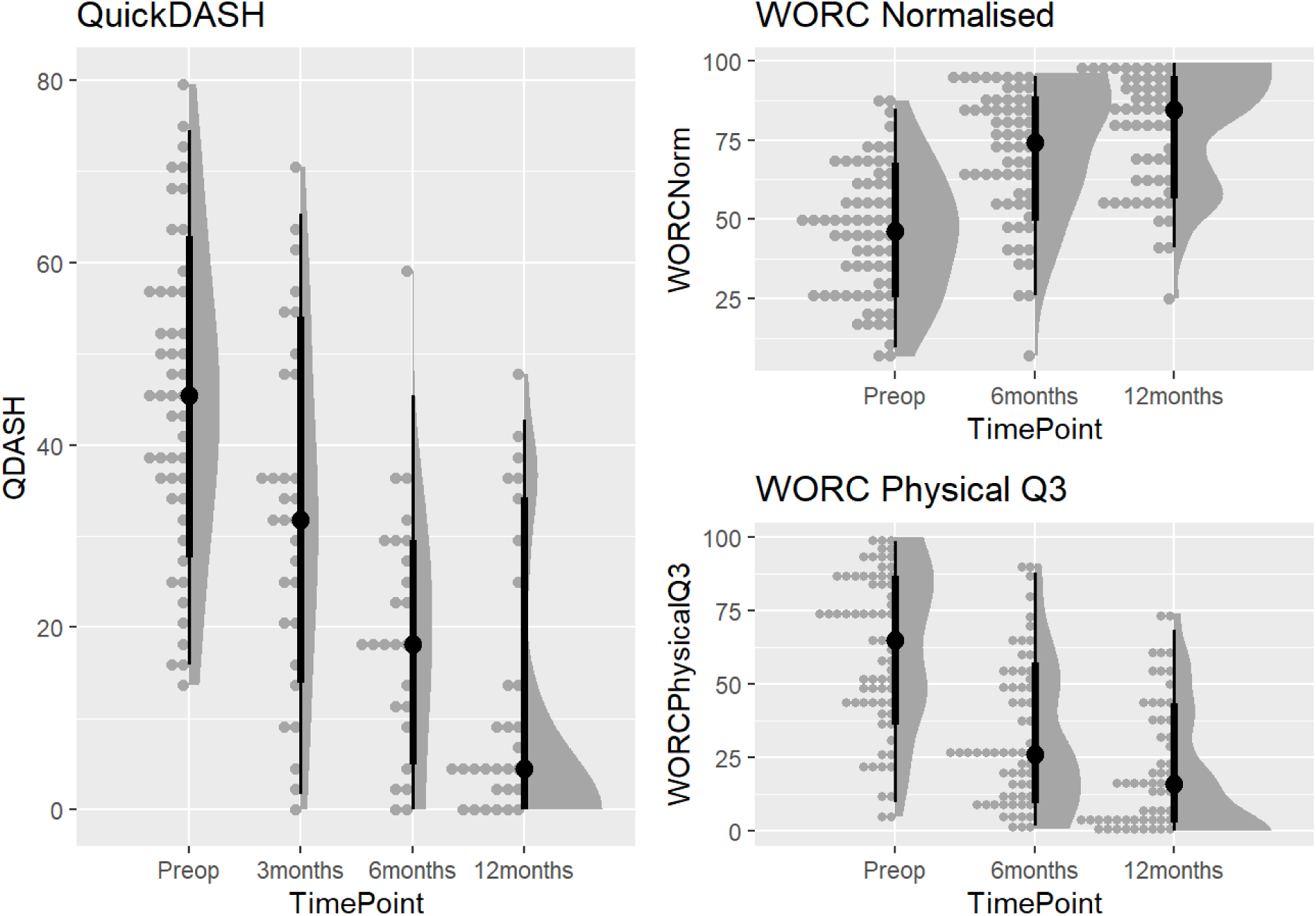
Change in distribution over time for adjusted and imputed QuickDASH, WORC Index and Q3 of the Physical subscale with median and interquartile ranges (black dot and solid line)

## Discussion

The aim of this paper was to present a stage 2a IDEAL evaluation of a third-generation biocomposite suture anchor in a registry sub-analysis of patients presenting with repairable rotator cuff tear for arthroscopic repair in a regional practice, describing the incidence of all-cause failures, adverse events and the trajectory of patient-reported outcomes up to 12 months follow up.

The key findings were a low incidence of postoperative complications and no anchor-related intraoperative events, with no requirement for reoperation or revision, as well as improvement in patient-reported outcomes aligned with expectations derived from the relevant literature The reported benefits of PLGA/β-TCP implants is improved absorption and increased osteoconductivity, with low rates of imaging-observed adverse events (Barber et al., 2017; Suroto et al., 2023). We report low-grade postoperative complications after ARCR using a PLGA/β-TCP anchor (Healix Advance BR). A registry-based analysis of ARCR (N = 1891) (Felsch et al., 2021) reported an overall incidence of all complication types of 18.5% (95%CI 16.6 - 20.4) at 6 months follow-up. The upper confidence limit of the present complication incidence (18.6%) aligns well to this estimate and may be improved with a larger sample. Overall, the use of the anchor of interest was not associated with a higher than expected rate of postoperative complications in this sample, with just one case of persistent post-surgical pain observed.

The clinical outcomes reported here cannot be readily compared with other studies reporting on the use of the anchor of interest (Chung et al., 2018; Di Gennaro et al., 2022) as they rely on postoperative MRI to assess imaging-based complications and did not report clinical presentations of known complications or adverse events. The presence of cysts is frequently reported, and varies widely at 12 month follow up, ranging from 10.8% (Ro et al., 2019) to 60% (Chung et al., 2018, 2019), but may not be clinically significant in the majority of cases (Chung et al., 2019). Further, one comparative study observed no relationship between perianchor fluid and the integrity of cuff repair, while the persistence of perianchor cysts postoperatively may be associated with a larger tear size and greater tendon retraction (Chung et al., 2019). Perianchor osteolysis (Pawaskar et al., 2015) and foreign body reactions (Nusselt et al., 2010) have been reported for biodegradable anchors (Park et al., 2017), but the presence of radiolucent rings around these anchors (indicating osteolysis) have not been correlated with healing/retear or clinical outcomes in rotator cuff repair (Park et al., 2017).

To the best of our knowledge, there is one other study that has assessed patient-reported outcomes with the use of PLGA implants, specifically the anchor of interest in our study (Di Gennaro et al., 2022). The questionnaires utilised are slightly different (QuickDASH vs DASH) although well correlated with each other, there are subtle differences in scores that can accumulate error to the point of a minimal clinically important difference (Gummesson et al., 2006). Also the baseline DASH reported in their study is higher, with the reported mean for the sample one point below the maximum score for the questionnaire and the between-patient standard deviation exceeding the maximum score. The demographics of the studies also differ, with Di Gennaro et al reporting half the proportion of males compared to the present cohort. The baseline QuickDASH in this study is more typical of what has been observed for patients undergoing rotator cuff repair (Macdermid et al., 2015; Shibahashi et al., 2024), with the minimally clinical important difference of 15.9 - 20 points observed by six months in this study (Macdermid et al., 2015). Average WORC Index scores for the present cohort were also comparable at baseline (albeit higher) and at 12 months follow up in a meta-analysis of rotator cuff repair comprising 15 studies (N = 1371) (Sahoo et al., 2021). Of note however, the results of the individual question asking about shoulder weakness revealed a proportion of patients experiencing weakness at 12 month follow up. Considering the lack of imaging-confirmed failures in this series, these results may relate to a subset of patients at risk of displaying signs of failure below the clinical threshold for seeking additional care. Worse PROMs and reduced strength are associated with retears in this population (Jancuska et al., 2018). Future work should target this subset of patients for imaging follow up to confirm cuff status.

### Limitations

The study is embedded within a clinical registry that is vulnerable to specific biases that should not be ignored (Scholes et al., 2023). Of particular relevance is selection bias, as the capacity for patients to engage in the process may be limited by language, technological and competency barriers. Thus the present cohort is not presented as representative of the entire cuff repair population, as evidenced by the majority male sample, above-average baseline scores, and private funded patient status. Future evaluations may include postoperative imaging to assess imaging-based complications that are particularly relevant for this type of anchor (Barber et al., 2017) and longer follow up. However, the outcome measures of this study are well validated and representative of the clinically relevant functional outcome status of the cohort. Further, judgements made about success or failure at this stage (2a) may not reflect the full spectrum of clinically relevant effects that could be observed with longer term follow-up (McCulloch et al., 2013). The alternative however, would be to wait for years for the result of definitive trials, which may not be possible without short-term observational studies as presented here.

## Conclusion

The findings of this IDEAL stage 2a evaluation at up to 12 months follow up reveal an acceptable incidence of adverse events, no requirement for reoperation and improvement of patient-reported outcomes in those treated for rotator cuff tear with ARCR with a third-generation biocomposite suture anchor by an experienced surgeon. These results should provide sufficient motivation for a larger evaluation or randomised trial to further evaluate efficacy in a bigger and more diverse population.

## Data Availability

All data produced in the present study are available upon reasonable request to the authors

## Acknowledgements

The authors would like to acknowledge the participation of the patients in the PRULO registry and contributions of the staff at Geelong Orthopaedics for the conduct of the study. The authors would also like to thank Milad Ebrahimi (EBM Analytics) and other team members for assistance with data collection and extraction for this study.

