## Supplementary 2 for "Stage 2a IDEAL evaluation of a third-generation biocomposite suture anchor in arthroscopic rotator cuff repair: Subgroup cohort analysis of the PRULO registry with 12-month follow up"

### Clinical outcome of Healix Advance BR (Biocryl Rapide)

Corey Scholes

23-Jul-2024

#### 1. Introduction

This analysis links to the manuscript of the Healix BR product as one of two companion publications assessing new-to-market hardware. The dataset is derived from the PRULO registry snapshot and live tables. A protocol has been previously prepared for the registry (Scholes et al. 2023).

##### 1.0.1 Preparation

Load up required packages in advance. Citations applied to each library at first use in the text.

1. Load required packages
2. Check if packages are installed, if not, install them

##### 1.0.2 Aim

To describe the clinical and patient-reported outcomes, in patients presenting for surgical review of shoulder pathology and electing to undergo reconstruction or repair of soft-tissue structures with a biodegradable anchor (Healix Advance BR, Depuy-Mitek, USA), at a private, regional orthopaedic clinic between 2020 - 2024.

##### 1.0.3 Hypothesis

No hypotheses have been constructed for this case series as yet.

### 2. Analysis Methods

#### 2.1 Reporting

The study was reported according to the RECORD guidelines (Benchimol et al. 2015) and companion checklist.

The study was reported according to the RECORD guidelines (Benchimol et al. 2015) and companion checklist. The analysis was conducted in RStudio IDE (RStudio v2024.04.1+748 "Chocolate Cosmos" Release ) and (v1.4.4) (Allaire and Dervieux 2024) using R R version 4.4.0 (2024-04-24 ucrt) and associated packages to perform the following;

- Data import and preparation
- Sample selection
- Describe missingness
- Data manipulation and analysis of;
  - Patient characteristics
  - Pathology characteristics
  - Management and surgical technique
  - Treatment and repair survival
  - Adverse events and complications
  - Patient reported outcomes

- QuickDASH
- WORC Index (Normalised)

##### 2.1.1 RECORD [4] - Study Design

Subgroup analysis of a clinical registry embedded into private practice. Observational, cohort design.

#### 2.2 Data Import and Preparation

Retrieve and format data from live tables and registry snapshot. Using *openxlsx* (v1.8) (Barbone and Garbuszus 2024) to retrieve static snapshot files and *googlesheets4* (v1.1.1) (Bryan 2023) to retrieve live database tables. Text and code output are integrated using the *epoxy* package (v1.0.0) (Aden-Buie 2023).

Read in live tables

Combine dataframes into one to conduct analysis using *tidyverse* (v2.0.0) (Wickham et al. 2019).

##### 2.2.1 RECORD [5] - Setting

The PRULO registry is based in a regional private practice for upper limb orthopaedics (Scholes et al. 2023).

The registry has 2679 treatment records with the first patient enrolled 13 October 2020 and the final treatment record created 22 March 2024.The registry snapshot was extracted on 26 March 2024. Patients are followed for up to 2 years after surgery to capture treatment outcomes and patient-reported outcome measures (PROMs).

#### 2.3 Record [6] Participants

##### 2.3.1 Record [6.1] Sample selection

Identify cases receiving the suture of interest. Cases were identified by SKUs identified from the SKU database maintained as part of implant tracking within the registry. Cases were not restricted by available follow up.

Inclusion criteria;

- Case involves anchor of interest
- Case is the index procedure within the registry (first use of suture)
- Patient has not withdrawn consent for inclusion of data in the registry
- Treatment record is eligible for surgery (it has occurred)

Data manipulation (add columns and filter tables based on column values) was performed with *tidyverse* and conversion to display format using *gt* (v0.11.0) (Iannone et al. 2024).

Table 1: Summary of SKUs (Reference) used to identify cases of interest from PRULO registry

| Size (mm) | Description | Category | Reference |
|---|---|---|---|
| 4.5 | Healix Advance BR OrthoCord | Anchor + Suture | 10886705021314 |
| 4.5 | Healix Advance BR 3 strand Orthocord | Anchor + Suture | 10886705021321 |
| 5.5 | Healix Advance BR OrthoCord | Anchor + Suture | 10886705021338 |
| 5.5 | Healix Advance BR 3 strand Orthocord | Anchor + Suture | 10886705021345 |
| 6.5 | Healix Advance BR OrthoCord | Anchor + Suture | 10886705021369 |
| 6.5 | Healix Advance BR 3 strand Orthocord | Anchor + Suture | 10886705021376 |
| 4.5 | Healix Advance BR PermaCord | Anchor + Suture | 10886705024803 |
| 4.5 | Healix Advance BR 3 strand PermaCord | Anchor + Suture | 10886705024735 |
| 5.5 | Healix Advance BR PermaCord | Anchor + Suture | 10886705024810 |
| 5.5 | Healix Advance BR 3 strand PermaCord | Anchor + Suture | 10886705024827 |
| 6.5 | Healix Advance BR PermaCord | Anchor + Suture | 10886705024834 |
| 6.5 | Healix Advance BR 3 strand PermaCord | Anchor + Suture | 10886705024681 |
| 4.5 | Healix Advance BR PermaTape (Blue) | Anchor + Tape | 10886705027798 |
| 4.5 | Healix Advance BR PermaTape (White/Blue) | Anchor + Tape | 10886705027804 |
| 5.5 | Healix Advance BR PermaTape (Blue) | Anchor + Tape | 10886705027811 |
| 5.5 | Healix Advance BR PermaTape (White/Blue) | Anchor + Tape | 10886705027828 |
| 6.5 | Healix Advance BR PermaTape (Blue) | Anchor + Tape | 10886705029266 |
| 6.5 | Healix Advance BR PermaTape (White/Blue) | Anchor + Tape | 10886705029273 |
| 4.5 | Healix Advance BR Dynacord (x2) with Needles | Anchor + Suture | 10886705029440 |
| 4.5 | Healix Advance BR Dynacord (x2) | Anchor + Suture | 10886705029402 |
| 4.5 | Healix Advance BR Dynacord (x3) | Anchor + Suture | 10886705029396 |
| 5.5 | Healix Advance BR Dynacord (x2) | Anchor + Suture | 10886705029464 |
| 5.5 | Healix Advance BR Dynacord (x3) | Anchor + Suture | 10886705029457 |
| 5.5 | Healix Advance BR Dynacord (x2) with Needles | Anchor + Suture | 10886705029471 |
| 6.5 | Healix Advance BR Dynacord (x2) | Anchor + Suture | 10886705029525 |
| 6.5 | Healix Advance BR Dynacord (x3) | Anchor + Suture | 10886705029518 |
| 6.5 | Healix Advance BR Dynacord (x2) with Needles | Anchor + Suture | 10886705029532 |

The registry snapshot was filtered to remove any non-index procedures from the master table.

Of the 69 records in the mastersheet, 0 treatment records had withdrawn consent for data inclusion and 1 had declined to participate in PROMs.

##### 2.3.2 Record [6.2] Algorithm validation

Record selection code was cross-checked by manual record checking within the registry snapshot for a subset (N = 10) of cases.

##### 2.3.3 Record [6.3] Data linkage

No data linkage was utilised for this analysis.

#### 2.4 Record [7] Variables

Key variables defined as part of this analysis are summarised in Table 2 below.

Table 2: Summary of key variable definitions in the analysis

| Category | Variable | Comments | Citation |
|---|---|---|---|
| Patient Characteristics | Insurance Status | Recode from account data from practice management system to insurance status | |
| Pathology | Primary diagnosis | Free text coded using ICD-10 international | |
| | CuffRetraction | Defined as per *modified* Patte grading | (Lädermann et al. 2016) |
| | CuffCondition | Fatty infiltration as assessed by Goutallier scale | (Fuchs et al. 1999) |
| | TearPattern | Shape the tear makes within the margins of the cuff as viewed in the transverse plane | (Lädermann et al. 2016) |
| | OtherShoulder Pathology | Free-text coded as present [Yes] or not [No] | |
| Management - Surgery | RepairAugment | Techniques used to augment the repair | |
| | CuffTension | Surgeon perceived tension to restore anatomical footprint of repair | |
| | RepairQuality | Surgeon subjective rating of the repair quality | |
| Survival | TreatmentStatu s | Labelled as failure after review of clinical notes indicating construct failure (non-operative management) OR reoperation involving removal of index repair hardware | |
| | RetearStatus | Adverse event involving image-confirmed retear or hardware loosening | |
| Adverse Events | Modidifed sink grade | Modification of the Sink grading of complication severity | (Felsch et al. 2021) |
| Patient-Reported | WORC Physical | How much weakness do you | (Kirkley, Alvarez, and |

| Category | Variable | Comments | Citation |
|---|---|---|---|
| Outcomes | Q3 | experience in your shoulder? | Griffin 2003) |

#### 2.5 Record [8] Data sources

Data was sourced directly from the PRULO clinical registry.

#### 2.6 Record [9] Bias

For a discussion of biases in the context of the clinical registry utilised for this analysis, refer to (Scholes et al. 2023). Specific to this analysis, the following considerations are noted;

Table 3: Biases in analysis of observational cohort of a clinical registry

| Bias | Definition | Source | Mitigation |
|---|---|---|---|
| Misclassification | Treatment record labelled into incorrect cohort. PROMs package not aligned to | (Benchimol et al. 2015) | Clinical notes reviewed by experienced reviewer and matched to ICD10 code by definition. |
| Confounder | An variable of interest and a target outcome simultaneously influenced by a third variable | (Tennant et al. 2020) | PROMs analysis incorporated adjustment for age and sex |
| Missing data | The absence of a data value where a treatment record is eligible to have a data value collected | (Carroll, Morris, and Keogh 2020) | Multiple imputation utilised |
| Prevalent user | Follow-up starts after eligible individuals have started the treatment. The follow-up time is left-truncated | (Nguyen et al. 2021) | Eligibility and enrollment is performed prior to treatment offering for any patient or new presentation. Index procedures identified for analysis are followed prior to surgery occurring. |
| Selection | Treatments are selected based on post-treatment criteria | (Nguyen et al. 2021) | Unable to be mitigated fully - records are identified by presence of hardware code associated with suture of interest |

| Bias | Definition | Source | Mitigation |
|---|---|---|---|
| Immortal time | Individuals need to meet eligibility criteria that can only be assessed after follow-up has started | (Nguyen et al. 2021) | Patients enrolled at time of diagnosis |
| Pseudoreplication | Analyse data while ignoring dependency between observations. Inadequate model specification. | (Davies and Gray 2015; Lazic 2010) | Cluster for patient in survival (all-cause failure and retear). Utilise mixed effects linear model (lme4::lmer) for PROMs analysis with treatment identifier as random effect |

#### 2.7 Record [10] Sample size

Sample size was derived based on the available records from the Registry at the time of analysis.

#### 2.8 Record [11] Quantitative variables

The anterior-posterior (AP) and mediolateral (ML) dimensions of the cuff tear were reported and multiplied to calculate tear area (mm^2). The tear was also classified according to (Rashid et al. 2017).

- **Small** tears were defined as full-thickness defects in the supraspinatus tendon under 1 cm in the anterior–posterior (AP) dimension.
- **Medium** tears were defined as full-thickness defects in the supraspinatus tendon only, greater than 1 cm and less than 3 cm in the AP dimension.
- **Large** tears involved full-thickness defects of both the supraspinatus and infraspinatus tendons, greater than 3 cm, and less than 5 cm in the AP dimension.
- **Massive** tears involved all 3 tendons (supraspinatus, infraspinatus, and subscapularis) and were greater than 5 cm in the AP dimension.

Partial tears were left labelled as *partial*. Ultimately recoded tear classification based on AP tear length, as the involvement of other tendons for tears of small length was not adequately defined in the original paper.

Data was read in from database table to determine account type.

Process data tables to summarise procedure and surgery details.

The complications table was processed for further analysis.

Complication severity grading was performed according to (Felsch et al. 2021).

Mastertable was prepared to analyse repair failure time to event.

Tables were rearranged to analyse patient-reported outcomes (PROMs).

Tables were modified to track anchor usage.

#### 2.9 Record [12] Statistical methods

A number of analytical techniques were employed to i) clean the data inputs as well as ii) evaluate missingness in the dataset and iii) complete the descriptive analysis of;

- Patient characteristics
- Pathology details
- Patient, implant and adverse event time to event
- Patient-reported outcomes

##### 2.9.1 Record [12.1] Access to population

The registry system represents all cases presenting to the rooms of a surgical group within Geelong, Australia using the implant of interest from the inception of the clinical registry to the analysis date. All reviewed charts from the operating surgeons practice records (electronic medical record) were entered into database and the present analysis draws data from a regular compilation of the registry records (snapshot) produced quarterly by the registry administration team.

##### 2.9.2 Record [12.2] Data cleaning methods

Diagnosis and complication description free text fields were pre-processed to remove relational terms (stopwords) and expand abbreviations to improve clarity.

Dates of events (preceding and subsequent surgical records; adverse events including mortality) relative to index surgery date were assessed using coded checks to flag anomalies and were resolved by further manual review to resolve inconsistencies or discrepancies with the chart review input data stored in the registry database.

##### 2.9.3 Record [12.3] Data linkage

Not applicable

##### 2.9.4 Record [12.4] Missingness evaluation and management

Missingness was assessed with visualisation and table functions in the *naniar* package (v1.1.0) (Tierney and Cook 2023) and compiled into figures using *patchwork* (v1.2.0.9000) (Pedersen 2023).

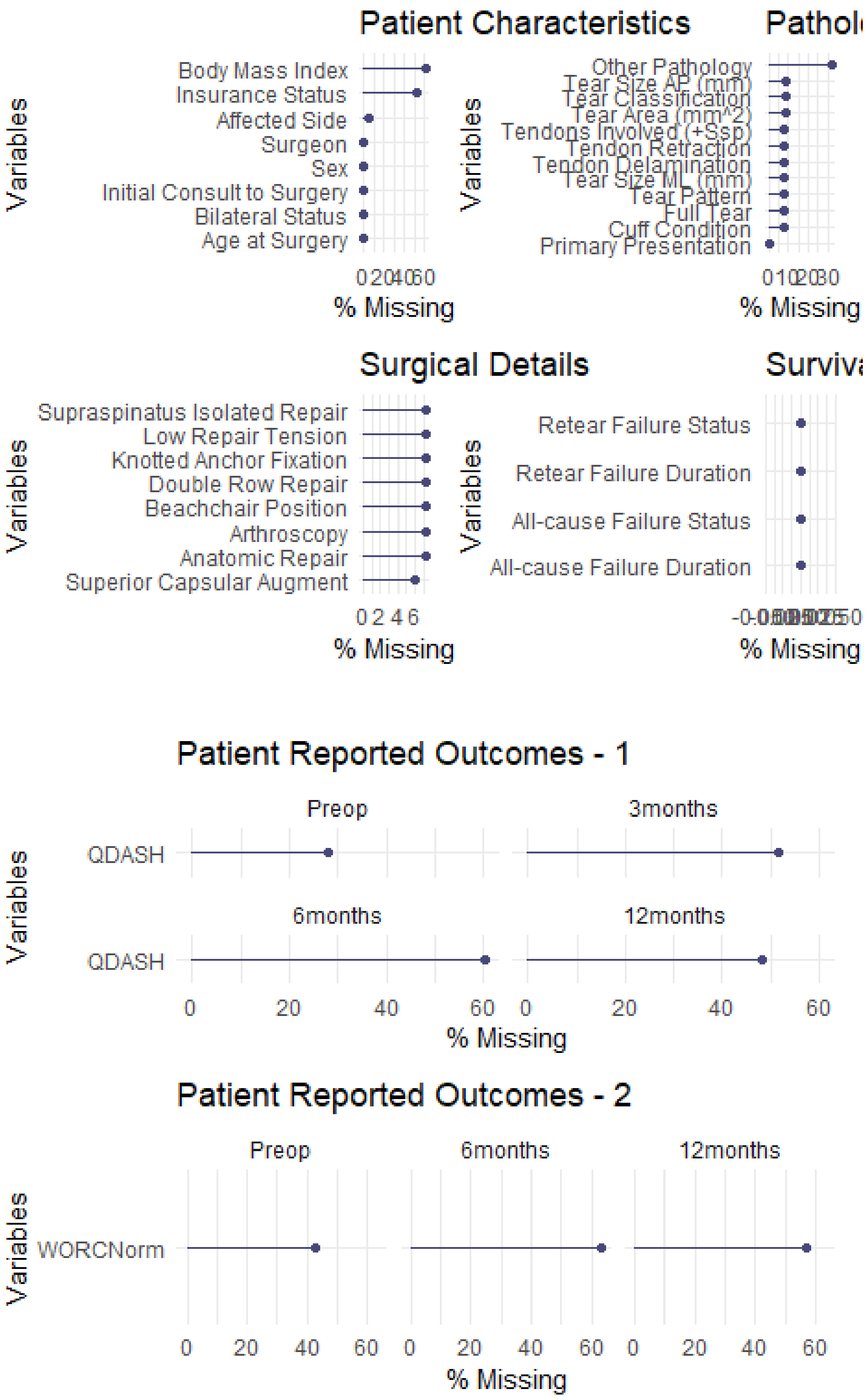

Figure 1: Missingness by variable

The compliance for the QuickDASH at baseline was 72.1% and for the WORCNorm it was 57.4%.

The compliance for the QuickDASH was 51.7% and for the WORCNorm it was 43.1% at 12months.

The data tables were sliced to the required columns (PROMs and adjunct columns) in preparation for multiple imputation using chained equations (White, Royston, and Wood 2010) with the *mice* package (v3.16.0) (Buuren and Groothuis-Oudshoorn 2011).

### 3. Analysis Results

#### 3.1 Record [13] Participants

The initial export from the registry returned 2679 records of all types.

##### 3.1.1 Record [13.1] Treatment selection

A flow chart of individual treatment episodes (treatments) was generated using the *consort* package (v1.2.2)(Dayim 2023) and prepared for display with the *knitr* package (v1.48) (Xie 2024).

The diagram below summarises recruitment and categorisation of patients into the PRULO registry.

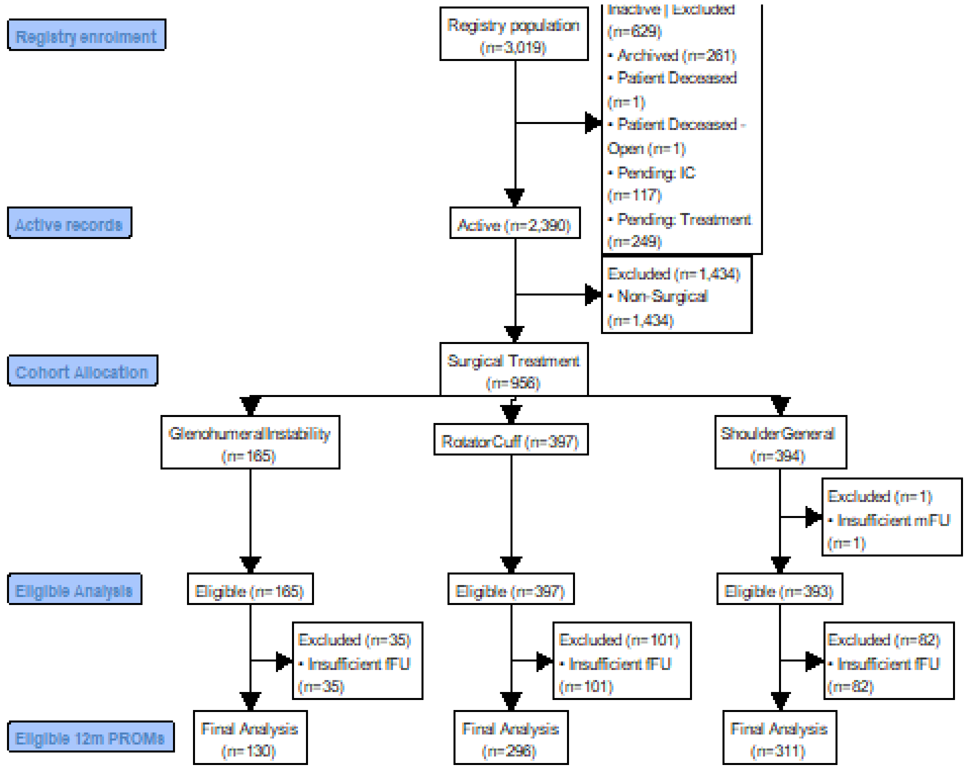

Figure 2: Flow chart of treatment record inclusion and separation into cohorts for analysis. mFU - Minimum follow up (30 days).

The table below summarises patient diagnoses in the PRULO registry.

Table 4: Summary of diagnoses by ICD-10 code

*Table 4: Summary of PRULO diagnoses by ICD-10 (top 5 by cohort*

| RegistryCohortName | ICD10 | n |
|---|---|---|
| GlenohumeralInstability | S43.42 | 61 |
| GlenohumeralInstability | S43.0 | 53 |
| GlenohumeralInstability | M25.31 | 31 |
| GlenohumeralInstability | M24.4 | 24 |
| GlenohumeralInstability | M24.21 | 4 |
| RotatorCuff | M75.1 | 261 |
| RotatorCuff | S46.0 | 107 |
| RotatorCuff | S43.43 | 26 |
| RotatorCuff | M75.3 | 18 |
| RotatorCuff | M75.4 | 10 |
| ShoulderGeneral | M19.0 | 220 |
| ShoulderGeneral | M75.0 | 47 |
| ShoulderGeneral | S42.0 | 34 |
| ShoulderGeneral | M12.0 | 25 |
| ShoulderGeneral | S43.5 | 20 |

#### 3.2 Record [14] Patient and record characteristics

Patient characteristics for cases receiving the anchor of interest are summarised in Table 5.

Table 5: Summary of patient characteristics

| **Characteristic** | **N** | **N = 68** | **95% CI**[1] |
|---|---|---|---|
| Age at Surgery, Mean (SD) | 68 | 61 (9) | 58 - 63 |
| Female, % (n) | 68 | 31% (21) | 21 - 43 |
| Non-dominant, % (n) | 65 | 40% (26) | 28 - 53 |
| Surgeon, % (n) | 68 | | |
| B | | 100% (68) | 93 - 100 |
| BMI, Mean (SD) | 26 | 23 (14) | 18 - 29 |

| **Characteristic** | **N** | **N = 68** | **95% CI**[1] |
|---|---|---|---|
| Bilateral, % (n) | 68 | 16% (11) | 8.7 - 28 |
| Exam to surgery delay (weeks), Mean (SD) | 68 | 10 (10) | 7.7 - 13 |
| Insurance Type, % (n) | 33 | | |
| DVA[2] | | 3.0% (1) | 0.16 - 18 |
| Private | | 88% (29) | 71 - 96 |
| TAC[3] | | 3.0% (1) | 0.16 - 18 |
| Uninsured | | 6.1% (2) | 1.1 - 22 |

[1]CI = Confidence Interval

[2]DVA = Department of Veterans Affairs

[3]TAC = Transport Accident Commission

##### 3.2.1 Record [14.1] Pathology characteristics

Pathology characteristics for cases receiving the anchor of interest are summarised in Table 6.

Table 6: Summary of pathology characteristics

| Characteristic | Available Sample | Summary Statistic | 95% CI[1] |
|---|---|---|---|
| Primary Presentation, % (n) | 68 | 100 (68) | 93 - 100 |
| Full Tear, % (n) | 63 | 98 (62) | 90 - 100 |
| Fatty Infiltration, % (n)[2] | 63 | | |
| 0[2] | | 11 (7) | 5.0 - 22 |
| 1[2] | | 51 (32) | 38 - 63 |
| 2[2] | | 33 (21) | 22 - 46 |
| 3[2] | | 4.8 (3) | 1.2 - 14 |
| Tendon Retraction, % (n)[3] | 63 | | |
| I[3] | | 33 (21) | 22 - 46 |
| II[3] | | 44 (28) | 32 - 57 |
| III[3] | | 19 (12) | 11 - 31 |

| Characteristic | Available Sample | Summary Statistic | 95% CI[1] |
|---|---|---|---|
| IV[3] | | 3.2 (2) | 0.55 - 12 |
| Tendon Delamination, % (n) | 63 | 71 (45) | 58 - 82 |
| Tendons Involved (+Supraspinatus), % (n) | 63 | | |
| Infraspinatus | | 22 (14) | 13 - 35 |
| Infraspinatus; Subscapularis | | 11 (7) | 5.0 - 22 |
| Infraspinatus; Teres Minor; Subscapularis | | 1.6 (1) | 0.08 - 9.7 |
| Subscapularis | | 13 (8) | 6.0 - 24 |
| Subscapularis (isolated) | | 16 (10) | 8.3 - 28 |
| Supraspinatus (isolated) | | 37 (23) | 25 - 50 |
| Tear Size AP (mm), Mean (SD) | 62 | 24 (11) | 21 - 27 |
| Tear Size ML (mm), Mean (SD) | 63 | 19 (9) | 17 - 22 |
| Tear Area (mm^2), Mean (SD) | 62 | 551 (588) | 402 - 700 |
| Tear Classification, % (n)[4] | 62 | | |
| Large[4] | | 16 (10) | 8.4 - 28 |
| Massive[4] | | 3.2 (2) | 0.56 - 12 |
| Medium[4] | | 76 (47) | 63 - 85 |
| Partial[4] | | 1.6 (1) | 0.08 - 9.8 |
| Small[4] | | 3.2 (2) | 0.56 - 12 |
| Tear Pattern, % (n) | 63 | | |
| Crescent | | 49 (31) | 37 - 62 |
| L | | 17 (11) | 9.4 - 30 |
| Reverse L | | 14 (9) | 7.1 - 26 |
| U | | 16 (10) | 8.3 - 28 |
| V | | 3.2 (2) | 0.55 - 12 |
| Other Pathology, % (n) | 46 | 43 (20) | 29 - 59 |

| Characteristic | Available Sample | Summary Statistic | 95% CI[1] |
|---|---|---|---|

[1]CI = Confidence Interval

[2]Fuchs et al 1999

[3]Modified Patte Grading (Lädermann et al., 2016)

[4](Rashid et al., 2017)

##### 3.2.2 Record [14.2] Management summary

Surgical details are summarised in Table 7.

Table 7: Summary of management and surgical details

| **Characteristic** | ****Available Sample**** | ****Summary Statistic**** | **95% CI**[1] |
|---|---|---|---|
| Arthroscopy, % (n) | 63 | 100% (63) | 93 - 100 |
| Beachchair Position, % (n) | 63 | 100% (63) | 93 - 100 |
| Supraspinatus (isolated) Repair, % (n) | 63 | 35% (22) | 24 - 48 |
| Double Row Repair, % (n) | 63 | 83% (52) | 70 - 91 |
| Knotted Anchor Fixation, % (n) | 63 | 100% (63) | 93 - 100 |
| Superior Capsular Augment, % (n) | 64 | 0% (0) | 0.00 - 7.1 |
| Low Repair Tension, % (n) | 63 | 67% (42) | 54 - 78 |
| Anatomic Repair, % (n) | 63 | 79% (50) | 67 - 88 |

[1]CI = Confidence Interval

##### 3.2.3 Record [14.3] Follow up

The sample overall had a median follow up of 101 months.

Table 8: Summary of case followup (months) for the sample included for analysis

| **Characteristic** | **No further followup**, N = 1[1] | **Ongoing**, N = 67[1] |
|---|---|---|
| TreatDuration | 20 (20, 20) | 102 (79, 139) |

[1]Median (IQR)

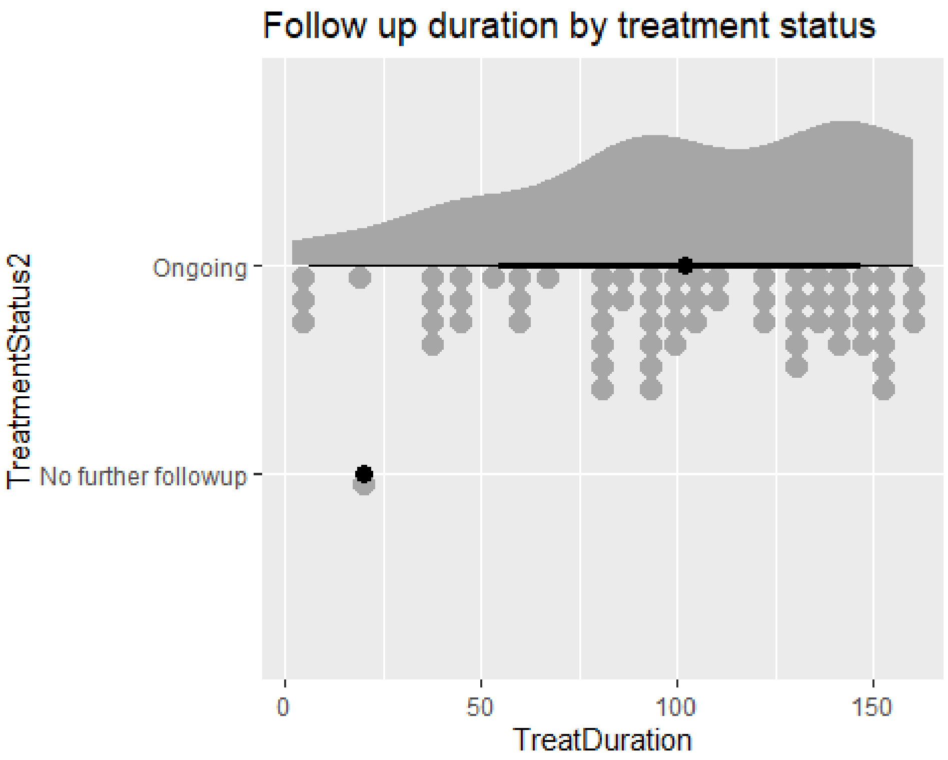

Figure 3: Summary of follow up duration for the included sample.

#### 3.3 Record [15] Outcomes

##### 3.3.1 Record [15.1] Treatment survival (free from all-cause failure)

There were no revisions or retears observed in the present sample.

##### 3.3.2 Record [15.2] Treatment survival (free from retear)

There were no retears observed in the present sample.

##### 3.3.3 Record [15.3] Adverse events and complications

Of the 68 cases included in the analysis, there were 6 treatments observed with 6 adverse events of any kind, equating to an incidence of 8.8 (95%CI, 3.6 - 18.9). There were 0 reoperations and 0 events observed intraoperatively.

A summary of the adverse events observed is included in Table 10.

Table 10: Summary of adverse events

| Characteristic | Available Sample | Summary Statistic |
|---|---|---|
| Complication Nature, % (n) | 6 | |
| Capsulitis - Stiffness | | 50% (3) |

| Characteristic | Available Sample | Summary Statistic |
|---|---|---|
| Pain - Other | | 50% (3) |
| Severity Grade, % (n) | 6 | |
| 1 | | 67% (4) |
| 2 | | 17% (1) |
| 4 | | 17% (1) |

##### 3.3.4 Record [15.4] Patient-reported outcome measures

The QuickDASH total score and WORC Normalised Index, as well as Question 3 of the Physical sub-scale of the WORC were visualised using the *ggdist* (v3.3.2) (Kay 2023) and *ggplot2* (v3.5.1) (Wickham 2016)packages. Plots were arranged using the *patchwork* package.

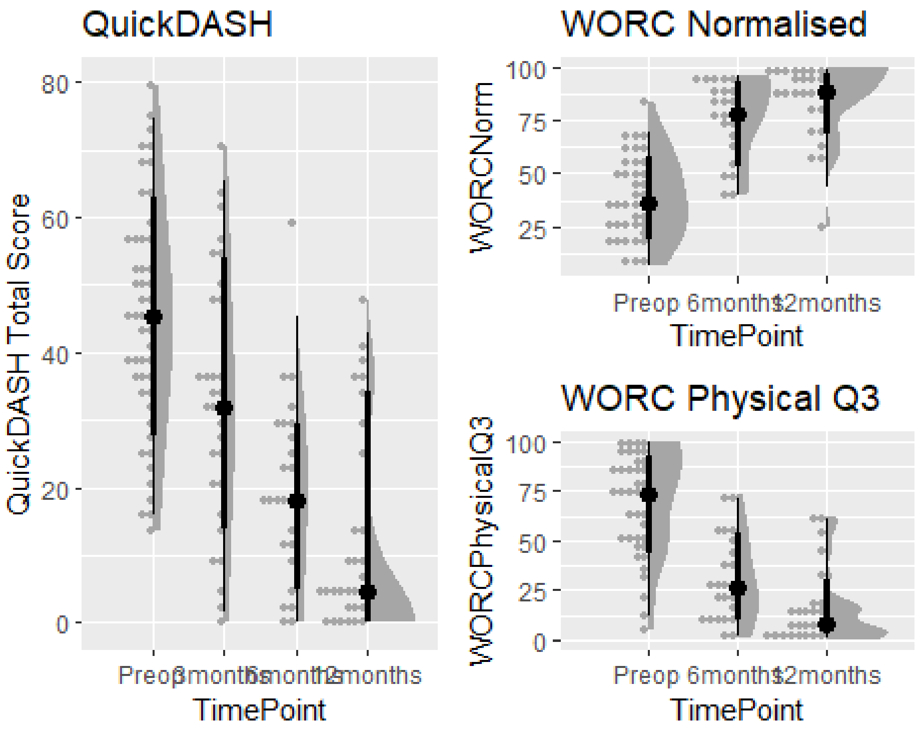

Figure 6: Summary of patient reported outcomes trajectories by timepoint

#### 3.4 Record [16] Main results

The imputed datasets for QDASH and WORC were modeled with a linear mixed effects model in *lme4* (v1.1.35.5) (Bates et al. 2015)and summarised with *broom.mixed* (v0.2.9.5) (Bolker and Robinson 2022). Up to a 38.7 point improvement in QuickDASH total score was observed (Table 11), as well as 47.1 and 54 point improvements in WORC Index Normalised and WORC Physical Question3 respectively (Table 12). Distributions of model-predicted results illustrated variability in recovery trajectories within all PROMs measures (Figure 7).

Table 11: Summary of pooled linear model results QDASH

| **Characteristic** | **Beta** | **95% CI**[1] | **p-value** |
|---|---|---|---|
| TimePoint | | | |
| Preop | — | — | |
| 3months | -12.11 | -18.28, -5.95 | <0.001 |
| 6months | -25.38 | -32.11, -18.65 | <0.001 |
| 12months | -32.11 | -38.50, -25.71 | <0.001 |
| Age at Surgery | -0.03 | -0.43, 0.37 | 0.873 |
| Male vs Female | -8.19 | -16.39, 0.00 | 0.050 |

[1]CI = Confidence Interval

Table 12: Summary of pooled linear model results WORC

| | Normalised Index | | | Physical Q3 | | |
|---|---|---|---|---|---|---|
| **Characteristic** | **Beta** | **95% CI**[1] | **p-value** | **Beta** | **95% CI**[1] | **p-value** |
| TimePoint | | | | | | |
| Preop | — | — | | — | — | |
| 6months | 31.81 | 18.54, 45.08 | <0.001 | -30.56 | -43.94, -17.18 | <0.001 |
| 12months | 38.69 | 28.21, 49.16 | <0.001 | -43.75 | -55.59, -31.91 | <0.001 |
| Age at Surgery | 0.28 | -0.18, 0.74 | 0.229 | -0.43 | -1.13, 0.26 | 0.204 |
| Male vs Female | 9.18 | -1.11, 19.47 | 0.078 | -8.84 | -21.47, 3.79 | 0.161 |

[1]CI = Confidence Interval

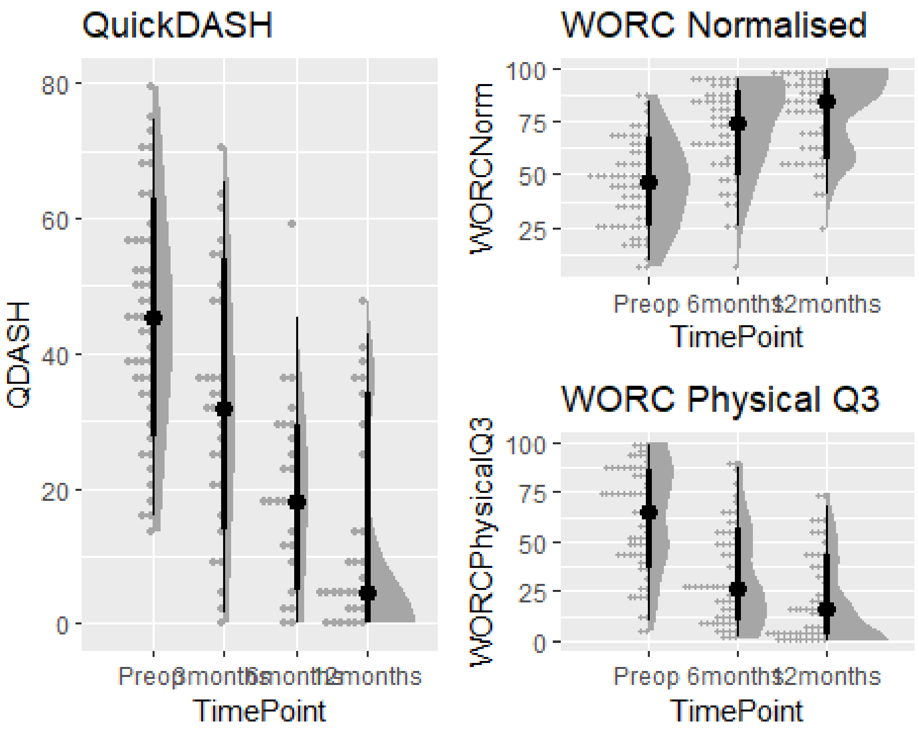

Figure 7: Model predicted PROMs trajectories across time points.

**Note:** RP feedback 2-Mar was to add % exceeding MCID for WORCNorm at 6m and 12m follow up. The MCID data for WORC in rotator cuff is highly volatile and requires a careful analysis in its own right to appropriately select and implement an MCID analysis in this context.

Table 13a: Summary of model-predicted QuickDASH by TimePoint

| **Characteristic** | **Preop**, N = 49[1] | **3months**, N = 31[1] | **6months**, N = 25[1] | **12months**, N = 30[1] |
|---|---|---|---|---|
| QuickDASH | 45 (34, 57) | 32 (20, 48) | 18 (11, 30) | 5 (2, 22) |

[1]Median (IQR)

Table 12b: Summary of model-predicted WORC by TimePoint

| **Characteristic** | **Preop**, N = 68[1] | **6months**, N = 63[1] | **12months**, N = 58[1] |
|---|---|---|---|
| WORCNorm | 46 (27, 60) | 74 (57, 87) | 85 (62, 93) |
| WORCPhysicalQ3 | 65 (43, 87) | 26 (12, 51) | 16 (5, 38) |

[1]Median (IQR)

#### 3.5 Record [17] Sensitivity analyses

Based on the distribution changes in QDASH and WORC over time, a sensitivity analysis was performed on the model structure using the complete case dataset. A comparison was made between quantile regression using the *quantreg* package (v5.98) (Koenker 2023)and an ordinary least squares linear model from the *rstats* package (v4.4.0) (2022) and a linear mixed effects model with the *lme4* package (v1.1.35.5) (Bates et al. 2015). Results were tabulated using the *modelsummary* package (v2.1.1) (Arel-Bundock 2022) as rq models are not supported in *gtsummary*.

Table 13: Comparison of linear model types to assess patient-reported outcomes

| | RQ | LM | ME |
|---|---|---|---|
| (Intercept) | 50.0 | 56.2 | 53.7 |
| | se = 13.5 | se = 10.7 | se = 12.8 |
| | [23.6, 76.4] | [35.0, 77.4] | [28.3, 79.2] |
| TimePointRecode3months | -11.4 | -11.7 | -12.1 |
| | se = 4.6 | se = 3.7 | se = 3.1 |
| | [-20.3, -2.4] | [-19.1, -4.3] | [-18.3, -5.9] |
| TimePointRecode6months | -25.0 | -24.6 | -25.4 |
| | se = 3.9 | se = 4.0 | se = 3.4 |
| | [-32.7, -17.3] | [-32.6, -16.7] | [-32.1, -18.6] |
| TimePointRecode12months | -38.6 | -31.8 | -32.1 |
| | se = 3.4 | se = 3.8 | se = 3.2 |
| | [-45.2, -32.0] | [-39.3, -24.4] | [-38.5, -25.7] |
| AgeAtTreatment | 0.0 | -0.1 | 0.0 |
| | se = 0.2 | se = 0.2 | se = 0.2 |
| | [-0.4, 0.4] | [-0.4, 0.2] | [-0.4, 0.4] |
| Sex2Male | -6.8 | -7.1 | -8.2 |
| | se = 3.8 | se = 3.4 | se = 4.1 |
| | [-14.3, 0.7] | [-13.7, -0.4] | [-16.4, 0.0] |
| SD (Intercept TreatmentUID) | | | 10.1 |
| | | | se = 1.8 |
| | | | [7.1, 14.3] |
| SD (Observations) | | | 12.9 |
| | | | se = 1.0 |
| | | | [11.0, 15.2] |
| Num.Obs. | 135 | 135 | 135 |
| R2 | 0.386 | 0.412 | |
| R2 Adj. | | 0.389 | |
| R2 Marg. | | | 0.406 |
| R2 Cond. | | | 0.630 |
| AIC | 1146.9 | 1143.7 | 1114.6 |
| BIC | 1164.3 | 1164.0 | 1137.9 |
| ICC | | | 0.4 |
| Log.Lik. | | -564.854 | |
| F | | 18.050 | |
| RMSE | 16.23 | 15.88 | 11.06 |

The comparison between models revealed an underestimate of the difference in 12month score to preoperative baseline of 7.7 points for the QuickDASH (14.7%) in the mixed effects linear model, compared to the quantile regression (50th percentile).

#### 4. Export Files

Export images files for reporting.

Monitoring).” *Ecology and Evolution* 5 (22): 5295–5304. https://doi.org/10.1002/ece3.1782.

Dayim, Alim. 2023. “Consort: Create Consort Diagram.” https://CRAN.R-project.org/package=consort.

Felsch, Quinten, Victoria Mai, Holger Durchholz, Matthias Flury, Maximilian Lenz, Carl Capellen, and Laurent Audigé. 2021. “Complications Within 6 Months After Arthroscopic Rotator Cuff Repair: Registry-Based Evaluation According to a Core Event Set and Severity Grading.” *Arthroscopy: The Journal of Arthroscopic & Related Surgery* 37 (1): 50–58. https://doi.org/10.1016/j.arthro.2020.08.010.

Fuchs, Bruno, Dominik Weishaupt, Marco Zanetti, Juerg Hodler, and Christian Gerber. 1999. “Fatty Degeneration of the Muscles of the Rotator Cuff: Assessment by Computed Tomography Versus Magnetic Resonance Imaging.” *Journal of Shoulder and Elbow Surgery* 8 (6): 599–605. https://doi.org/10.1016/s1058-2746(99)90097-6.

Iannone, Richard, Joe Cheng, Barret Schloerke, Ellis Hughes, Alexandra Lauer, and JooYoung Seo. 2024. “Gt: Easily Create Presentation-Ready Display Tables.” https://CRAN.R-project.org/package=gt.

Kay, Matthew. 2023. “Ggdist: Visualizations of Distributions and Uncertainty.” https://doi.org/10.5281/zenodo.3879620.

Kirkley, Alexandra, Christine Alvarez, and Sharon Griffin. 2003. “The Development and Evaluation of a Disease-Specific Quality-of-Life Questionnaire for Disorders of the Rotator Cuff: The Western Ontario Rotator Cuff Index.” *Clinical Journal of Sport Medicine* 13 (2): 84–92. https://doi.org/10.1097/00042752-200303000-00004.

Koenker, Roger. 2023. “Quantreg: Quantile Regression.” https://CRAN.R-project.org/package=quantreg.

Lädermann, Alexandre, Stephen S. Burkhart, Pierre Hoffmeyer, Lionel Neyton, Philippe Collin, Evan Yates, and Patrick J. Denard. 2016. “Classification of Full-Thickness Rotator Cuff Lesions: A Review.” *EFORT Open Reviews* 1 (12): 420–30. https://doi.org/10.1302/2058-5241.1.160005.

Lazic, Stanley E. 2010. “The Problem of Pseudoreplication in Neuroscientific Studies: Is It Affecting Your Analysis?” *BMC Neuroscience* 11 (1). https://doi.org/10.1186/1471-2202-11-5.

Nguyen, Van Thu, Mishelle Engleton, Mauricia Davison, Philippe Ravaud, Raphael Porcher, and Isabelle Boutron. 2021. “Risk of Bias in Observational Studies Using Routinely Collected Data of Comparative Effectiveness Research: A Meta-Research Study.” *BMC Medicine* 19 (1). https://doi.org/10.1186/s12916-021-02151-w.

Pedersen, Thomas Lin. 2023. “Patchwork: The Composer of Plots.” https://CRAN.R-project.org/package=patchwork.

R Core Team. 2022. "R: A Language and Environment for Statistical Computing." https://www.R-project.org/.

Rashid, Mustafa S, Cushla Cooper, Jonathan Cook, David Cooper, Stephanie G Dakin, Sarah Snelling, and Andrew J Carr. 2017. "Increasing Age and Tear Size Reduce Rotator Cuff Repair Healing Rate at 1 Year." *Acta Orthopaedica* 88 (6): 606–11. https://doi.org/10.1080/17453674.2017.1370844.

Scholes, Corey, Kevin Eng, Meredith Harrison-Brown, Milad Ebrahimi, Graeme Brown, Stephen Gill, and Richard Page. 2023. "Patient Registry of Upper Limb Outcomes (PRULO): A Protocol for an Orthopaedic Clinical Quality Registry to Monitor Treatment Outcomes." *Journal of Surgical Protocols and Research Methodologies* 2023 (4). https://doi.org/10.1093/jsprm/snad014.

Tennant, Peter W G, Eleanor J Murray, Kellyn F Arnold, Laurie Berrie, Matthew P Fox, Sarah C Gadd, Wendy J Harrison, et al. 2020. "Use of Directed Acyclic Graphs (DAGs) to Identify Confounders in Applied Health Research: Review and Recommendations." *International Journal of Epidemiology* 50 (2): 620–32. https://doi.org/10.1093/ije/dyaa213.

Tierney, Nicholas, and Dianne Cook. 2023. "Expanding Tidy Data Principles to Facilitate Missing Data Exploration, Visualization and Assessment of Imputations" 105. https://doi.org/10.18637/jss.v105.i07.

White, Ian R., Patrick Royston, and Angela M. Wood. 2010. "Multiple Imputation Using Chained Equations: Issues and Guidance for Practice." *Statistics in Medicine* 30 (4): 377–99. https://doi.org/10.1002/sim.4067.

Wickham, Hadley. 2016. "Ggplot2: Elegant Graphics for Data Analysis." https://ggplot2.tidyverse.org.

Wickham, Hadley, Mara Averick, Jennifer Bryan, Winston Chang, Lucy D'Agostino McGowan, Romain François, Garrett Grolemund, et al. 2019. "Welcome to the Tidyverse" 4: 1686. https://doi.org/10.21105/joss.01686.

Xie, Yihui. 2024. "Knitr: A General-Purpose Package for Dynamic Report Generation in r." https://yihui.org/knitr/.
